# Brain Alterations in COVID Recovered Revealed by Susceptibility-Weighted Magnetic Resonance Imaging

**DOI:** 10.1101/2022.11.21.22282600

**Authors:** Sapna S Mishra, Rakibul Hafiz, Rohit Misra, Tapan K. Gandhi, Alok Prasad, Vidur Mahajan, Bharat B. Biswal

**Affiliations:** Department of Electrical Engineering, Indian Institute of Technology Delhi, Hauz Khas, New Delhi, 110016, India; Department of Biomedical Engineering, New Jersey Institute of Technology (NJIT), 323 Dr Martin Luther King Jr Blvd, Newark, 07102, NJ, USA; Irene Hospital, Haryana, India; Mahajan Imaging Center, Hauz Khas, New Delhi, India

**Keywords:** COVID-19, SARS-CoV-2, Post-COVID symptoms, Long COVID, Susceptibility Weighted Imaging, Fatigue

## Abstract

The increasing number of reports of mild to severe psychological, behavioral, and cognitive sequelae in COVID-19 survivors motivates a need for a thorough assessment of the neurological effects of the disease. In this regard, we have conducted a neuroimaging study to understand the neurotropic behavior of the coronavirus. We hypothesize that the COVID recovered subjects have developed alterations in the brain which can be measured through susceptibility differences in various regions of brain when compared to healthy controls (HCs). Hence we performed our investigations on susceptibility weighted imaging (SWI) volumes. Fatigue, being of the most common symptoms of Long COVID has also been studied in this work. SWI volumes of 46 COVID and 30 HCs were included in this study. The COVID patients were imaged within six months of their recovery. We performed unpaired two-sample t-test over the pre-processed SWI volumes of both the groups and multiple linear regression was performed to observe group differences and correlation of fatigue with SWI values. The group analysis showed that COVID recovered subjects had significantly higher susceptibility imaging values in regions of the frontal lobe and the brain stem. The clusters obtained in the frontal lobe primarily show differences in the white matter regions. The COVID group also demonstrated significantly higher fatigue levels than the HC group. The regression analysis on the COVID group yielded clusters in anterior cingulate gyrus and midbrain which exhibited negative correlations with fatigue scores. This study suggests an association of Long COVID with prolonged effects on the brain and also indicates the viability of SWI modality for analysis of post-COVID symptoms.

**Highlights:**

- Susceptibility weighted imaging is used for neuroimaging study of Long COVID.
- A group-level study is performed to analyze the effects of COVID on the brain.
- COVID survivors showed susceptibility differences in the frontal lobe and brainstem.
- Analyzed the relationship between MRI data of COVID survivors and fatigue scores.

## 1. Introduction

Coronaviruses (CoV) are a large family of viruses that cause illnesses ranging from the common cold to more severe diseases such as middle east respiratory syndrome (MERS) and severe acute respiratory syndrome (SARS). A novel coronavirus (nCoV), named severe acute respiratory syndrome coronavirus 2 (SARS-CoV-2), is a strain of coronavirus recently found in humans. In December 2019, the first case of coronavirus disease 2019 (COVID-19) was reported. Since then, the virus has caused a global pandemic claiming over 6.3 million lives worldwide as of June 2022 [1]. Most of these deaths are consequences of the pulmonary complications caused by the coronavirus infection. However, there has been an increase in reports of the virus attacking the human central nervous system (CNS) [2]. Neurological symptoms and signs like confusion, delirium, headache, loss of memory, and disorder of consciousness have been observed and reported worldwide [3]. Apart from the direct impact of the CNS by the virus, the reported neurological manifestations might be related to pathophysiological mechanisms of para-infections or post-infections. Moreover, newer studies report the neurological manifestations of coronavirus in those who survive the virus infection [4]. This situation is worrisome considering the large population affected by this pandemic, including over 524 million people worldwide and 42 million in India [1]. The increasing number of reports of mild to severe psychological, behavioral, and cognitive sequelae in the survivors motivates a need for an adequate and thorough assessment of the neurological effects of COVID-19. A neuroimaging-based study to understand the post-COVID effects, also known as Long COVID, can aid the research community to determine the neurotropic behavior of coronavirus and detect the brain regions being affected directly or indirectly by the virus.

Susceptibility weighted imaging (SWI) combines the phase and the magnitude information obtained from magnetic resonance imaging [5]. The three-dimensional high-resolution gradient echo (GRE) sequence used in the SWI modality enables it to utilize the phase data. When combined in a specific manner with these phase data the magnitude data yields SWI volumes. This fully flow-compensated MR imaging technique employs long echo time and short flip angles. It exploits magnitude susceptibility differences of various compounds, such as blood, paramagnetic iron, and diamagnetic calcium, present in different brain tissue types [6]. Thus SWI technique is useful in detecting several pathologies and conditions such as microhemorrhages, cerebral microbleeds, vascular malformations, trauma, and multiple sclerosis [5, 7, 8, 9].

Neurological effects of COVID-19 can be inferred from several symptoms, including loss of smell and taste, brain fog, and headaches [3]. Some people have complained about debilitating fatigue, loss of memory, and difficulty concentrating. In early research on the effects of COVID-19 on the brain, in-depth examinations were carried out on human brain tissues of patients who died after COVID contamination [10, 11]. A study by Lee and colleagues [10] found signs of inflammation and unusually bright and dark spots in the brain’s olfactory bulb, but it also reports no presence of SARS-CoV-2 in the brain. On the other hand, the work by Song and colleagues [11] suggests that the virus may directly infect the human CNS as the authors detected the presence of the virus in the brain’s cerebral cortex. In both these studies, the number of subjects is small. Hence, we could not make conclusive deductions about whether the coronavirus directly affects the brain or the pathologies present are secondary effects of the physiological complications caused by the virus in the human body. Later, to diagnose the abnormalities in living patients’ brains, the research community used neuroimaging techniques to further investigate the preliminary findings of deformities.

A descriptive review in by Gulko and colleagues has reported on multi-sequence MRI scans of 126 patients across seven countries and established the neurological pathologies caused by COVID-19, as observed in their surveyed research works [12]. This review includes observations of acute infarct, posterior reversible encephalopathy syndrome (PRES), leukoencephalopathy, cortical abnormality, and microhemorrhages in the different regions of the brain. Kremer and colleagues [13] conducted a retrospective study on 64 subjects’ neuroimaging data collected over multiple centers. This study reports several neurological aberrations in the accumulated data of COVID patients, with over 56% of the cohort having abnormalities. The significant pathologies seen by the authors were ischemic strokes and encephalitis. In other studies, loss of gray matter in the frontal lobe [14], arterial wall thickening [15],and inflammatory vascular pathologies [16] have also been reported in the MRI scans. In the reports of [17], researchers reported abnormal findings in brain computed tomography (CT)/MRI scans of 40% of the 35 COVID-19 patients during their hospitalization. Most common pathologies observed were microbleedings and restricted diffusion lesions. On the other hand, the arterial spin labeling (ASL) based study in [18] was carried out on 39 non-hospitalized COVID-19 recovered adults and compared with 11 healthy controls. This work revealed decreased cerebral blood flow (CBF) in the thalamus, orbitofrontal cortex, and basal ganglia.

The multi-sequence investigation by Griffanti and colleagues [19] showed lower gray matter density in frontal gyrus, lower gray matter metric in hippocampus, left superior division of the lateral occipital cortex from the structural imaging. The SWI exploration of the same study reported changes in the thalamus, and left hippocampus. In the SWI study of COVID patients on mechanical ventilation [20], Conklin and colleagues observed abnormal susceptibility signals in 11/16 patients whereas >10 lesions were reported in 50% of the subjects. These lesions were primarily identified in the corpus callosum region, subcortical, and deep white matter, with variable inclusion of the cerebellum and brainstem. In the case report of 68 year old male hospitalized with a diagnosis of coronavirus (SARS-CoV-2), hypointense areas were observed in bilateral thalamus, in the genu of the corpus callosum, and in the parietal juxtacortical white matter [21]. Another SWI study reported leukoencephalopathy and microbleeds in patients with COVID-19 [22]. To the best of our knowledge, no work on SWI based group-level analysis of COVID recovered patients has been reported in literature.

In this study, we attempt to understand the effects of COVID on the human brain using the SWI modality of MRI. The literature reports ischemic microvascular diseases occurring in COVID patients [22], which makes SWI an adequate modality for the analysis. However, there are limited literature reporting group-level results on the SWI analysis of COVID patients compared with respect to healthy controls (HCs). We started by recruiting a sample of patients who had been tested positive for the PCR test for COVID-19. We scanned these patients within six months of subsequent PCR negative status. We hypothesize that the COVID recovered subjects will develop alterations in the brain’s anatomy, which can be measured through susceptibility differences in various regions of brain, when compared to HCs. We expect that these surviving COVID-negative patients would have altered susceptibility values of brain tissues, compared to the HC group. Further we anticipate to observe a significant correlation between altered brain regions (gray or white matter) of COVID recovered subjects and self-reported fatigue scores. SWI modality of MR imaging can provide such information and enable us to test our hypothesis.

## 2. Materials and Methods

### 2.1. Participants

In this study MRI volumes of 46 (15 females) COVID-19 recovered patients, termed as COVID group in the scope of this work, and 30 (8 females) HCs were included with mean age of 34.67 ± 9.51 years for COVID group and 35.09 ± 11.37 years for HCs. These participants were recruited by the Indian Institute of Technology (IIT), Delhi, India, where they were imaged following all Institutional Review Board (IRB) guidelines and all patients gave informed consent prior to providing any behavioral or physical data.

### 2.2. Clinical Assessment

We have clinical details of 38 out 46 patients of the COVID group during their COVID-19 infection period and we also managed to gather post-COVID symptoms of 29 (12 females; mean age = 32.62 ± 8.19 years) out of the 46 patients. These 29 volunteers and 19 (5 females; mean age = 30.63 ± 9.79 years) random samples from the HC cohort also filled-up a form of fatigue analysis where they were asked to rate the effects of fatigue in different spheres (work, education, personal and social) of their daily lives from 0 to 5.

Most common symptoms during COVID-19 were reported to be fever (33/38), cough (26/38) and body ache (25/38). Loss of sense of smell (16/38) and taste (12/38) were also highly observed symptoms along with difficulty in breathing (20/38). The severity of these manifestations of COVID-19 varied among the subjects recruited for our scans. We had 18 patients who had been hospitalized and had severe COVID-19 whereas 5 of them reported moderate and the remaining 15 of them suffered from mild symptoms among the 38 patients whose details were made available to us.

On the other hand, 29 of 46 scanned patients who agreed to share their post-COVID symptoms showed a range of post-COVID symptoms. After negative RT-PCR test, these subjects reported to suffer from frequent body ache (achy muscle: 55.17% and achy joints: 44.83%), fatigue (72.41%), headache (44.83%) and hair loss (37.93%). Persistent breathing issues (13.8%) and bowel irritation (24.14%) were also detailed by some of the subjects. On the other hand, some specific complaints of lack of attention (48.27%), issues with memory (31.03%), and lack of sleep (41.38%) were common among these 29 patients. As mentioned earlier, these patients along with 19 HCs filled up a form to mention how the fatigue has affected different spheres of their respective lives: work, study, social and personal life. In this study, we mainly focused on the impact on the work sphere, where the average fatigue score in the sub-set of the COVID group was found to be 2.93/5 with standard deviation of 1.067 and that of the HC group was 0.63 ± 0.76.

### 2.3. Imaging

#### 2.3.1. Anatomical MRI

T1-weighted images were acquired using a 3T GE scanner in 3D imaging mode with a fast BRAVO sequence. The scanner had a 32 channel head coil. The imaging parameters were inversion time (TI) = 450 ms; Flip angle = 12°, field of view (FOV) = 256 mm × 256 mm, number of slices = 152 (sagittal), slice thickness = 1.00 mm and spatial isotropic resolution of 1 mm.

#### 2.3.2. Susceptibility-Weighted MRI

SWI scans were acquired using the same scanner in 3D imaging mode with a fast SWAN sequence. The imaging parameters were, echo time (TE) = 25.0 ms and low repetition time(TR); Flip angle = 15° and FOV = 224 mm × 224 mm; number of slices = 256, 256, and 196 in axial, coronal and sagittal plane respectively; slice thickness = 1.4 mm.

### 2.4. Data Pre-processing

For the SWI sequence, we have used FSL (https://fsl.fmrib.ox.ac.uk/fsl) software for data pre-processing. After dicom to nifti conversion, the brain extraction tool (BET) of FSL was used for skull stripping of SWI and anatomical brain images. The skull-stripped T1-weighted volumes were co-registered to the SWI images using inter-modality registration of FLIRT algorithm [23]. These co-registered T1-weighted anatomical images were then warped to the MNI reference template and the transformation matrices thus obtained were utilized to register the SWI images to the MNI space, resulting in a cascaded spatial registration of the individual image. Since, susceptibility-weighted images are not absolute values but relative in nature, we normalize them before carrying out the analysis. We assumed a constant SWI value of the cerebrospinal fluid and normalized all registered individual images to the “water” value of the left and the right lateral ventricles [24], by using the Harvard-Oxford Subcortical Structural Atlas template [25].

**Table 1.** Group level statistics on participant demographics in HC and COVID group. The included demographics are age, sex and fatigue scores (for 29 COVID and 19 HC subjects). Keys: *p* = p-value, *stat* = test statistics, *t* = two-sample t-test statistic, *χ* = Chi-Squared statistic, *τ* = Wilcoxon Rank Sum test score, M = Male, F = Female, HC: Healthy Controls

| Measures | $p$ | $stat$ | HC, mean (SD) | COVID, mean (SD) |
| --- | --- | --- | --- | --- |
| Age (years) | 0.8673 | -0.1677 ($t$) | 34.667 (9.50) | 35.087 (11.37) |
| Sex | 0.5815 | 0.3038 ($\chi$) | 22M (8F) | 31M (15F) |
| Fatigue | 1.178e-07 | 5.297 ($\tau$) | 0.632 (0.76) | 2.93 (1.067) |

### 2.5. Statistical Analysis

To determine the differences in participants’ demographics, we performed two sample t-test on age and chi-squared test for sex differences between the two groups. For voxel-wise assessment of COVID-19 vs HC on SWI modality, we performed unpaired two-sample t-test over the pre-processed volumes of COVID and HC groups. Significant clusters were identified and the main effect of interest from the corresponding contrast maps representing the difference in mean beta scores from two groups was obtained by thresholding the t-score map values that survived the corrected threshold. To account for confounding effects that may explain some of the variances in the data, age and sex were also added as covariates of no interest. Cluster-based thresholding was applied at a height threshold of *p*_*unc*_ < 0.01, with family-wise error (FWE) correction at *p*_*F WE*_ < 0.05 for multiple comparisons. The cluster extent threshold (*k*_*E*_) obtained from this step was used to generate corrected statistical maps for the contrasts with significant effects. The clusters obtained after surviving FWE for extent threshold, *k*_*E*_ = 506 have been illustrated in Figure 1 with their respective cluster sizes and peak intensities.

**Figure 1:**
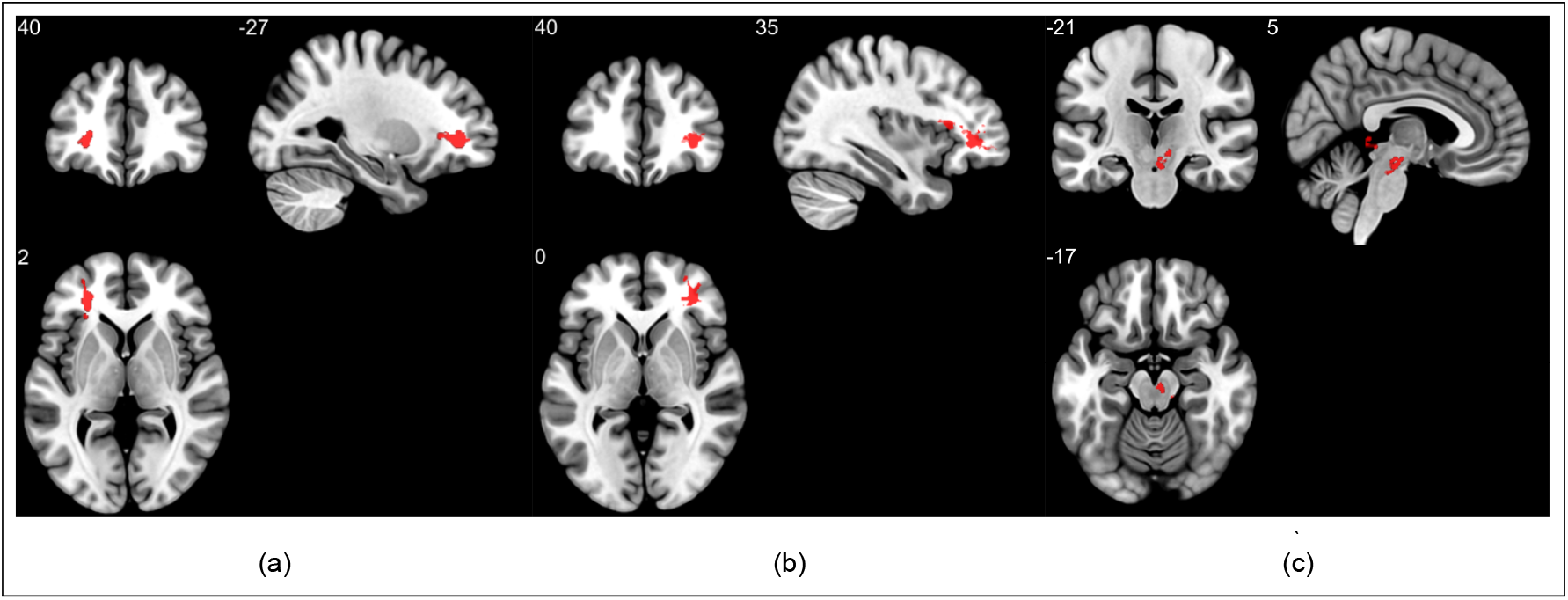
Group analysis on susceptibility weighted imaging exhibiting higher SWI values (lower susceptibilities) in the COVID group when compared to healthy controls. Three significant clusters were found primarily in the white matter regions of pre-frontal cortex and in the brainstem. The clusters (a) and (b) are observed bilaterally in the cerebral white matter near the orbito-frontal gyrus whereas (c) lies in the midbrain region.

In the fatigue study, the relationship of fatigue scores (in the work-sphere) with the COVID-19 recovered patients as was analyzed. Initially, the reported scores of the COVID and HC cohorts were compared and a significant difference in fatigue levels was observed between the two groups. Since the fatigue scores deviated from normality (Shapiro Wilk: *p* < 0.05), we used Wilcoxon’s rank-sum test to assess the group difference between HCs and COVID-19 subjects. Subsequently, to evaluate the correlation of fatigue score in work-sphere with the susceptibility values of the subset of COVID group, we performed a multiple linear regression analysis. Here, the voxel-wise SWI value was considered the response variable, the fatigue score was regarded as the covariate of interest, and sex and age were the confounding variables. Regions with a significant correlation between susceptibility values and fatigue score were identified using cluster-based thresholding at height threshold *p*_*unc*_ < 0.01 and FWE corrected at *p*_*F W E*_ < 0.05, for multiple comparisons.

## 3. Results

Group analysis on susceptibility-weighted images highlighted three major clusters in COVID group when compared to HCs (Figure 1). The first two clusters (Figure 1 (a) and (b)) were observed bilaterally in the white matter near the orbitofrontal gyri and the gray/white matter junctions. The left cluster had a volume of 711 voxels and a peak value of 4.0341, while the right cluster covered 1586 voxels and had a maximum *t*-value of 4.0913. The clusters bilaterally covered the uncinate fasciculus tract and anterior parts of the inferior fronto-occipital fasciculus tracts of white matter (JHU White-Matter Tractography Atlas). In addition, the third cluster was observed in the midbrain region of the brain stem with a volume of 506 voxels and maximum intensity of 3.8679.

In the fatigue study, the COVID group demonstrated significantly higher fatigue levels compared to HC group (using Wilcoxon’s rank-sum test: *τ* = 5.297, *p* = 1.178e-07). To evaluate the correlation of fatigue scores with SWI values, multiple linear regression analysis yielded four significant clusters in COVID-19 patients when compared with HCs (Figure 2). The cluster shown in Figure 2 (a), with a size of 1637 voxels, showed a peak intensity of 5.9 in a region covering the forceps minor tract, the thalamic radiations, and the anterior cingulate and paracingulate gyrus on the left side. The second cluster (Figure 2 (b)) was also observed in the left extension of the forceps minor tract in white matter areas whereas the peak intensity is observed in the left superior medial frontal gyrus. It covered 469 voxels and had a peak intensity of 5.844. Figure 2 (c) highlights a cluster of 595 voxels in the region encompassing midbrain and the left thalamus with peak intensity of 4.745. Further, the fourth cluster was again observed in a region covering the left anterior cingulate gyrus, the medial orbitofrontal cortex, and the subcallosal gyrus (Figure 2 (d)). The cluster had a volume of 1040 voxels and a peak intensity of 4.4573. Moreover, the SWI values in all the above clusters exhibited negative correlation with the fatigue scores in work-sphere (Spearman rank-order correlation coefficient, *ρ* ∈ [−0.6014, −0.647]).

**Figure 2:**
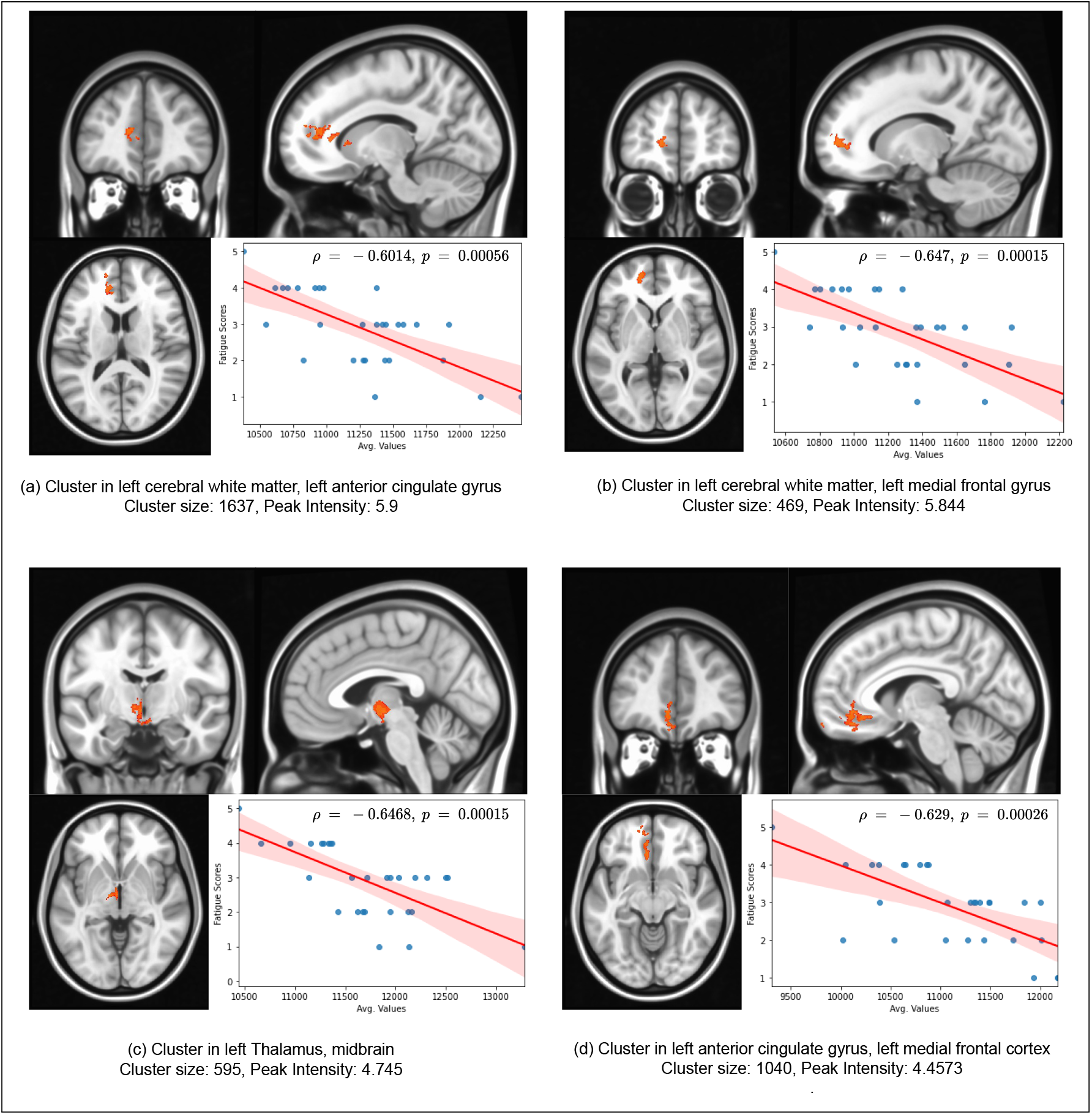
Results of group-level Susceptibility Weighted Imaging (SWI) analysis demonstrating significantly negatively correlated clusters with self-reported fatigue scores of work-sphere across the COVID recovered group. Here, *ρ* stands for the Spearman rank-order correlation coefficient. The blue colored dots represent the COVID recovered patients. The Avg. values in the x-axis denote the residuals plus the mean SWI values of the cluster across subjects added back after linear regression. The linear plot (red) represents the least squares regression line, and the shaded pink area depicts the 95% confidence interval.

**Table 2.** Lists all significant clusters in SWI signal intensity between COVID and HC groups. Significant FWE corrected differences were observed for COVID versus HC in WM and brainstem regions, while the inverse comparison of HC versus COVID yielded no supra-threshold clusters. Keys: HC: Healthy Controls, FWE: Family-wise Error, WM: White Matter.

| Cluster Index | Number of Voxels | Peak Intensity | Peak MNI Coordinates |  |  | COG MNI Coordinates |  |  | Anatomic Location |
| --- | --- | --- | --- | --- | --- | --- | --- | --- | --- |
|  |  |  | X | Y | Z | X | Y | Z |  |
| 1 | 711 | 4.034 | -28 | 29 | 3 | -28 | 43 | 2 | Left Cerebral White Matter in frontal lobe |
| 2 | 1586 | 4.09 | 34 | 42 | -1 | 30 | 40 | 3 | Right Cerebral White Matter in frontal lobe |
| 3 | 506 | 3.87 | 3 | -19 | -15 | 7 | -28 | -12 | Midbrain (Brainstem) |

**Table 3.** Information related to the negatively correlated clusters of SWI with fatigue scores of work-sphere across the 29 subjects of COVID recovered group. Significant clusters were obtained in the frontal lobe, anterior cingulate cortex and brainstem. All four cluster showed negative correlation with the fatigue scores. No cluster was found with positive correlation.

| Cluster Index | Number of Voxels | Peak Intensity | Peak MNI Coordinates |  |  | COG MNI Coordinates |  |  | Anatomic Location |
| --- | --- | --- | --- | --- | --- | --- | --- | --- | --- |
|  |  |  | X | Y | Z | X | Y | Z |  |
| 1 | 1637 | 5.953 | -12 | 41 | 14 | -13 | 34 | 9 | Left Anterior Cingulate Cortex |
| 2 | 469 | 5.844 | -14 | 51 | 2 | -14 | 53 | 4 | Left Superior Medial Frontal Gyrus |
| 3 | 595 | 4.74 | -3 | -8 | -5 | -4 | -9 | -5 | Midbrain |
| 4 | 1040 | 4.46 | -6 | 41 | -10 | -8 | 43 | -11 | Medial Orbitofrontal Gyrus |

## 4. Discussions

The ability of SWI modality to detect abnormalities in the brain has been widely reported in literature, where it has been demonstrated as a useful imaging technique for detecting microvascular pathologies and cerebral ischemia in white matter [26] and gray matter [27] regions. This motivates the use of SWI in investigating the effect of COVID-19 on the brain. In this study, the susceptibility weighted MR images of 46 COVID-19 recovered subjects and 30 healthy controls were acquired and group differences in susceptibility in the brain were studied. In the COVID-19 recovered cohort, nature of COVID infection ranged from mild to severe as inferred from self-reported symptoms. In the group comparison study, we have found significant clusters of abnormal susceptibility in the brain stem as well as in the white matter regions of the inferior frontal lobe which extend to the cortical gray/white matter junctions. These clusters with high SWI values may be a result of microvascular pathologies, cerebral ischemia, or other edematous pathophysiological in the brain. Similar decreased susceptibility in the brain has been reportedly associated with mild cognitive impairment [24] and post traumatic epilepsy [28].

Susceptibility weighted imaging has been used to investigate the effect of acute COVID-19 infection on the brain. Conklin and colleagues [20] have used SWI and presented their observations based on case-reports of 16 patients who were admitted to the intensive care unit with severe COVID-19 and showed neurological deficits. The investigation has revealed cerebral microvascular lesions in deep white matter (9/16 patients), cortical regions (9/16 patients) and the brainstem (3/16 patients). Moreover, a study conducted in France by Helms and colleagues [29] on COVID-19 patients admitted in the ICU with acute respiratory distress syndrome has also reported white matter micro-haemorrhages in the frontal lobe using SWI. On similar lines, the clusters found in our group based study also suggest abnormalities in corresponding regions of the brain. While, both these discussed studies have reported their observations on critically-ill COVID-infected patients, we have conducted our analysis on patients who have recovered from different levels of severity of COVID-19 infection.

The present study has shown significant bilateral SWI hyperintensities in the frontal, more specifically, orbitofrontal region, damage to which has consistently been reported in other studies on COVID patients as well [30]. Structural analysis between COVID affected and healthy brains by [31] has shown reduced cortical thickness and tissue contrast in the orbito-frontal gyrus. A perfusion study [32] using ASL has reported decreased cerebral blood flow in the orbital and medial frontal cortices. Multiple studies on COVID patients using positron emission tomography (PET)/CT imaging have also consistently reported hypometabolism of Fludeoxyglucose (FDG) in the frontal lobe [33, 34, 35, 36]. Abnormalities in the midbrain region of the brainstem highlighted in this study have also been reported in multiple studies on COVID patients [37]. In critically ill patients, there have been reports of cerebral microhemorrhages in the brain stem [38]. These may be secondary effects of COVID caused by severe hypoxia in these patients. In another study [39], neuroinflammatory changes were observed in the brainstem while investigating neuropathological features of the COVID affected brains. A study on COVID patients using FLAIR imaging reported 4 out of 6 patients showing abnormalities in the brain stem [40]. A study on a 62 year old COVID patient using PET imaging also revealed hypometabolism of FDG in the brain stem area [41], while another study employing PET reported increased voxel weights in the same region [36]. Consistent reports of COVID-19 affecting similar regions of the brain strengthens the evidence of neurotropic behavior of the virus.

It was observed that several symptoms reported in COVID and Long COVID patients such as loss of smell and taste, insomnia, fatigue, cognitive problems, and mental health issues could be attributed to the regions of abnormal susceptibilities in our study. While the frontal lobe is generally associated with cognitive control functions, influencing attention and memory [42], the orbitofrontal gyrus is essentially the secondary cortex for taste and smell processing [43]. It is connected directly to the primary olfactory and taste cortices. The orbitofrontal cortex is also associated with regulation of mood and emotions. On the other hand, the inferior frontal gyrus is linked to cognitive control over memory [44]. The clusters found in the frontal lobe are primarily located in the WM areas, which overlap with the uncinate fasciculus tract. This tract connects the limbic and paralimbic regions with the orbito-frontal gyrus [45]. Functional and structural abnormalities in these regions have been consistently reported in studies on COVID patients [46]. The limbic, paralimbic and orbito-frontal regions play a central role in emotion processing. The decreased connectivity amongst these regions may be linked to mental health issues and reduced memory [47]. Susceptibility differences were also observed in the brainstem, which is vital for cardio-respiratory regulation in the body, and injuries to this area are long lasting in nature [48]. Specifically, abnormalities in the midbrain and pons have been connected to multiple symptoms of Long COVID like fatigue, insomnia, anxiety, depression, headaches, and cognitive problems [49]. These observations are notable and consolidate our findings in relation to the behavioral and cognitive manifestations of COVID-19. They also supplement our understanding of the nature of neurotropism exhibited by the virus.

In the fatigue study, four clusters were identified in the brain that showed a significant negative correlation of mean SWI values of the clusters with self-reported fatigue scores of work-sphere. These correlations can be interpreted as positive correlations of the score with average susceptibilities of the clusters [28]. The obtained Spearman rank-order correlation coefficients for these clusters ranged from -0.647 to -0.601, with *p* < 0.001 for all the clusters (refer to Figure 2). Similar to our study, areas in the brain have been found to be related to fatigue in other reports as well. A functional connectivity analysis [50] has reported a negative correlation between fatigue and connectivity in the precuneus network which involves the left superior parietal lobule, the superior occipital gyrus, the angular gyrus, and precuneus. A volumetric study [51] on the brain of COVID affected patients has reported fatigue to be positively correlated to volume of the left posterior cingulate cortex, precuneus, and the superior parietal lobule. Another study [18] on blood perfusion in non-hospitalized COVID patients has reported increased CBF in superior occipital and parietal regions along with decreased perfusion in the inferior occipital regions.

The neural correlates of fatigue have been studied extensively using neuroimaging methods [52]. Fatigue has been shown to be associated with higher functional connectivity between regions, including the bilateral superior frontal gyri, the anterior cingulate gyrus, precuneus, right angular gyrus and posterior central gyrus, supplementary motor area, posterior cingulate gyrus and the thalamus [53]. In patients with myalgic encephalomyelitis (ME)/chronic fatigue syndrome (CFS), studies using single-photon emission computerized tomography (SPECT) [54] have reported a significantly higher perfusion in the anterior cingulate region along with reduced perfusion in other areas. The thalamus has been known to show significant changes in association with fatigue. In addition to the thalamus, the midbrain, cingulate, amygdala, pons, and the hippocampus have been reported to show inflammation in cases of fatigue [55]. In older people, the thalamus has been reported to exhibit neural correlates of perceived physical and mental fatigability [56]. These regions show substantial coherence with the gray and white matter regions identified to be correlated with fatigue in this study.

In this study, we have identified multiple clusters in gray matter as well as white matter brain regions of COVID patients where average susceptibility values are positively correlated (or average SWI values values are negatively correlated) with self-reported fatigue scores. The anterior cingulate cortex (ACC) is commonly known to be responsible for emotions and attention processing. The dorsal parts of the ACC contribute to cognitive functions while the ventral regions are involved in emotional processing [57]. The role of medial pre-frontal cortex in memory consolidation and retrieval of long term memory is also established in literature [58]. In studies on patients with depression, there have been reports of abnormal metabolic activity in the subcallosal gyrus [59]. This region was also identified in our study and may be related to emotional processing issues reported in Long COVID. The forceps minor tract and the anterior thalamic radiations connect the right and left frontal lobes. These neural circuits play a vital role in attention control and damage to these tracts has been associated with vascular cognitive impairments [60]. It must be observed that the established roles of the regions identified in this study have strong agreement with commonly reported symptoms of Long COVID.

Using high resolution T1 weighted anatomical images from the same subjects, we have also reported significant gray matter volume alterations in multiple brain regions from the limbic system and basal ganglia obtained using Voxel-Based Morphometry (VBM) analysis in [51]. The results from the present report complements the findings of the aforementioned conventional T1-weighted MR study. Both these studies together provide a holistic view into the neurological sequelae of COVID-19 as they utilize different mechanisms to investigate distinct properties of the brain. They suggest that the SARS-CoV-2 virus has structural and compositional effects on the brain. These neuroimaging impacts and their behavioral manifestations are visible even after months of recovery from the infection.

In conclusion, our results demonstrate group-level effects in COVID recovered patients, showing susceptibility differences in the GM and WM regions of the frontal lobe and brainstem. These observations are consistent with results reported in single-patient case studies carried out on SWI volumes and group studies using other MRI modalities. The present research will help the community to understand the impact of the SARS-CoV-2 virus on the human brain and hence propose some rehabilitative measures. In this study, we have also identified multiple clusters in gray matter, and white matter brain regions of COVID recovered patients where the average SWI values are negatively correlated with the self-reported fatigue scores. The roles of the clusters identified in this study have substantial agreement with commonly reported behavioral manifestations of post-COVID-19 fatigue. These brain regions have also been associated with fatigue in other neurodegenerative diseases. These deductions also indicate the viability of SWI modality for analysis of post-COVID symptoms. It also suggests an association of Long COVID with prolonged effects on the brain.

## 5. Limitations

While this study on the neurological basis of post-COVID symptoms using SWI has revealed significant abnormalities in susceptibilities of brain regions, there remain certain limitations that may be addressed to improve the impact and reliability of the study. Firstly, our study presents a group based analysis with a limited number of subjects which limits its transferability owing to unpredictable individual effects of each patient. Further, the application of SWI modality in clinical settings is slowly gaining relevance which hinders the immediate utility of our inferences for postulating the neuroimaging characteristics of post-COVID symptoms.

## Data Availability

Anonymized data used in the present study are available upon reasonable request to the authors.

## CRediT authorship contribution statement

**Sapna S Mishra:** Investigation, Methodology, Software, Formal Analysis, Resources, Data Curation, Writing - Original draft preparation, review and editing. **Rakibul Hafiz:** Investigation, Formal Analysis, Data Curation, Writing - Review and editing. **Rohit Misra:** Investigation, Formal Analysis, Writing - Review and editing. **Tapan K. Gandhi:** Conceptualization, Investigation, Resources, Supervision, Writing - Review and editing. **Alok Prasad:** Data Curation, Writing - Review and editing. **Vidur Mahajan:** Writing - Review and editing. **Bharat B. Biswal:** Conceptualization, Resources, Project Administration, Supervision, Writing - Review and editing.

## Acknowledgements

The work is supported by MeitY (Government of India) under grant 4(16)/2019-ITEA and Cadence Chair Professor fund awarded to Dr. Tapan Kumar Gandhi

